# Suppressed IgG4 class switching in dupilumab- and TNF inhibitor-treated patients after repeated SARS-CoV-2 mRNA vaccination

**DOI:** 10.1101/2023.09.29.23296354

**Authors:** Anika M. Valk, Jim B.D. Keijser, Koos P.J. van Dam, Eileen W. Stalman, Luuk Wieske, Maurice Steenhuis, Laura Y.L. Kummer, Phyllis I. Spuls, Marcel W. Bekkenk, Annelie H. Musters, Nicoline F. Post, Angela L. Bosma, Barbara Horváth, DirkJan Hijnen, Corine R.G. Schreurs, Zoé L.E. van Kempen, Joep Killestein, Adriaan G. Volkers, Sander W. Tas, Laura Boekel, Gerrit J. Wolbink, Sofie Keijzer, Ninotska I.L. Derksen, Melanie van Deelen, Gerard van Mierlo, Taco W. Kuijpers, Filip Eftimov, S. Marieke van Ham, Anja ten Brinke, Theo Rispens, T2B! Immunity against SARS-CoV-2 study group

## Abstract

**Background:** Repeated mRNA vaccination against SARS-CoV-2 has been shown to induce class switching to IgG4, a non-inflammatory human antibody subclass linked to tolerance. Although poorly understood, prolonged antigenic stimulation and IL-4 signalling may be instrumental in IgG4 switching. We and others have previously shown that widely used immunosuppressive drugs such as methotrexate (MTX) and TNF inhibitors (TNFi) have a minor inhibitory impact on humoral SARS-CoV-2 mRNA vaccination responses. However, the impact of such immunosuppressive drugs on IgG4 switching is unknown.

**Aim:** To study the impact of widely used immunosuppressive drugs (TNFi, MTX, or the IL-4 receptor-blocking antibody dupilumab on IgG4 skewing upon repeated SARS-CoV-2 mRNA vaccination.

**Methods:** Antibody responses to the receptor-binding domain (RBD) of the spike protein upon repeated SARS-CoV-2 mRNA vaccination were measured in 604 individuals including patients with immune-mediated inflammatory diseases treated with TNFi and/or MTX, or dupilumab, as well as healthy controls and untreated patients.

**Results:** We observed a substantial increase in the proportion of RBD-specific IgG4 antibodies (median 21%) in healthy/untreated controls after a third mRNA vaccination. This IgG4 skewing was absent when primary vaccination was adenoviral vector-based and was profoundly reduced in both dupilumab- and TNFi-treated patients (<1%), but only moderately in patients treated with MTX (7%).

**Conclusion:** Our results imply a major role for both IL-4/IL-13 as well as TNF in IgG4 class switching. These novel findings advance our understanding of IgG4 class switch dynamics, and may benefit future mRNA vaccine strategies, humoral tolerance induction, as well as treatment of IgG4 pathologies.

## Introduction

Repeated mRNA vaccination has been shown to induce class switching to IgG4 within the severe acute respiratory syndrome coronavirus 2 (SARS-CoV-2) specific B cell pool (1–3). The extent to which IgG4 class switching will affect immune protection to SARS-CoV-2 remains unclear, yet most likely depends on the mode of action of antibodies in SARS-CoV-2 infection and vaccination. In contrast to the proinflammatory IgG1 and IgG3 antibodies that dominate the initial responses, IgG4 has a more non-inflammatory character, with reduced affinity to most FcγRs and C1q, and therefore a limited potential for antibody dependent cellular cytotoxicity (ADCC) and complement-mediated effector functions (4,5). Furthermore, IgG4 is uniquely able to undergo Fab-arm exchange, resulting in a bispecific antibody that is functionally monovalent and unable to form immune complexes, further reducing its effector functions (6–9). However, IgG4 is generally associated with high levels of somatic hypermutation (SHM) and high affinity antigen binding, thereby contributing to efficient neutralization.

Both protective and pathogenic roles have been attributed to antibodies during the course of SARS-CoV-2 infection and vaccination. One mechanism of protection involves neutralization of the pathogen by interfering with the interaction of the spike protein to ACE2. In general, neutralising antibody titers induced by SARS-CoV-2 infection or mRNA vaccination correlate well with protection from infection (10–12). Furthermore, effector functions mediated by the Fc tail of IgG have been suggested to contribute to protection via complement-dependent or FcγR-dependent viral clearance (13,14). On the other hand, Fc-mediated effector functions might also contribute to excessive inflammation, leading to a more severe disease course. High levels of proinflammatory afucosylated antibodies were for instance found in patients admitted to the intensive care unit (ICU) following SARS-CoV-2 infection, suggesting a potential pathogenic role for the latter (15,16). Whether or not the weak potential of IgG4 to induce Fc effector functions is advantageous overall remains to be determined.

Another question is what is driving the switch towards non-inflammatory IgG4. Although key drivers of IgG4 class switching remain to be elucidated, prolonged antigenic stimulation and signalling along the T helper 2 axis have been shown to play a role (17–19). The repetitive nature of the SARS-CoV-2 mRNA vaccination strategy may therefore contribute to IgG4 class switching. While IgG4 class switching is a relatively uncommon event upon vaccinations, there are other examples of vaccine-induced antigen-specific IgG4 induction. The nature of the primary antigen challenge appears crucial in these cases. For instance, during HIV vaccination trials IgG4 switching was exclusively seen in response to bivalent recombinant protein-based vaccines (VAX003/VAX004 trails), but not after viral vector-based primary immunization (20,21). Furthermore, in case of pertussis vaccination, the acellular vaccine induced a stronger IgG4 switch in comparison to a whole cell vaccine variant (22,23). On the other hand, repeated tetanus toxoid immunization to produce hyperimmune anti-tetanus serum results in an IgG1-dominated response (24), illustrating that repeated vaccination in itself does not necessarily induce IgG4 class switching. Mechanistically, IgG4 switching has been associated with T helper 2 (Th2) responses, and IL-4 has been identified as an important cytokine for IgG switching in general and IgG4 in particular (25–31). Other cytokines that have been associated with (selective) IgG4 induction include IL-10 and IL-13, but details remain as yet unclear (5).

A potential concern with vaccinations is the response in patients treated with immunosuppressive medications. These drugs target different routes of the immune system and potentially interfere with vaccination strategies to activate the immune system. We and others have previously shown that the impact thereof varies greatly. Widely used drugs such as methotrexate (MTX) and the TNF inhibitors (TNFi) appear to have an overall limited impact on the immune response to SARS-CoV-2 mRNA vaccines, yet show slightly diminished antibody titers (32–34). Whether or not such immunosuppressive drugs may further enhance or prevent switching to the non-inflammatory IgG4 isotype is unknown. Specific drugs, in particular dupilumab that interferes with IL-4 signaling, may be expected to influence IgG4 switching. An allergen-specific IgG4 increase was nevertheless reported with long-term dupilumab use in atopic patients (35). In this study, we investigated the IgG4 switch of the anti-spike antibody response upon repeated mRNA vaccination in a large cohort of individuals, including patients treated with several widely used immunosuppressive drugs: dupilumab (IL-4R blocking antibody), TNFi, MTX, or a combination of TNFi and MTX.

## Methods

### Participants and study design

This study, part of the previously described Target-2-B! Immunity against SARS-CoV-2 vaccination cohort (32) included immune mediated inflammatory disease (IMID) patients treated with MTX, TNFi, the combination thereof, or dupilumab. Patients had no history of oncological or hematological disorders. Control groups consisting of IMID patients that were not treated with systemic immunosuppressants (disease controls, DC) and healthy controls (HC). Participants received two homologous doses of BNT162b2, mRNA-1273 or ChAdOx1 nCoV-19, followed by a booster dose of BNT162b2 or mRNA-1273. Serum samples were collected 28 days after each vaccination at-home fingerprick. Participants who experienced a SARS-CoV-2 infection prior to or during the study period were excluded from this sub-study. This study was approved by the medical ethical committee of the Amsterdam UMC (2020.194; trial registry NL74974.018.20 and EudraCT 2021-001102-30). All participants provided written informed consent.

### Anti-RBD ELISAs

To detect total IgG directed against the receptor binding domain (RBD) of the spike (S) protein of SARS-CoV-2 (Wuhan-Hu-1) in human serum, we performed a previously described in-house developed direct enzyme-linked immunosorbent assay (ELISA) (36,37). In short, this assay was calibrated using a pooled plasma standard obtained from convalescent healthy donors in May 2020, which was set at 100 arbitrary units (AU)/mL. The lower limit of quantification for samples tested at 1:1200 dilution was 1 AU/mL, with a >99% specificity cutoff determined at 4 AU/mL (36). This format was further adapted into an IgG4-specific ELISA. 96-well half-area microplates (Corning) with a working volume of 50 μL were coated overnight at 4°C with 1 μg/mL RBD in phosphate buffered saline (PBS; Fresenius Kabi). Plates were washed 5 times with PBS + 0.02% v/v Tween-20 (Merck, Germany) using an ELx405 ELISA washer (Biotek Instruments). Serum samples were diluted 1:200 in PBS + 0.1% v/v Tween-20 + 2 g/L gelatin (Merck, Germany)(PTG), added to the wells and incubated for 1 hour at RT. After washing, 0.5 μg/mL anti-human IgG4-HRP (MH164-4-HRP, Sanquin) in PTG was added and incubated for 30 minutes at RT. After one more washing step, 1-step Ultra TMB substrate (Thermo Scientific) diluted with milli-Q water in a ratio of 3:1 was added to wells. Reactions were stopped after approximately 7 minutes with an additional 50 μL 0.2M H_2_SO_4_ and optical density (OD) was measured at 450 nm and 540 nm with a Synergy 2 microplate reader (Biotek instruments). As a calibrator, the previously described COVA1-18 anti-RBD clone (38) was engineered with a human IgG4 heavy chain, analogously as described previously for IgG1 and IgG3 (36). The calibrator was twofold serially diluted starting from 0.025 μg/mL with a PTG blank. When tested in this manner in the total IgG assay, the IgG4 monoclonal demonstrated good linearity of dilution and parallelism with the 2020 pooled plasma standard starting from 0.25 AU/mL (1:400; Figure S1A). We thus calculated that 1 AU of 2020 pooled plasma produced similar IgG detection signal to 0.11 μg of IgG4 monoclonal, and IgG4 measurements could be expressed in AU/mL equivalent for comparison to total IgG measurements. Lastly, we determined a lower limit of quantification of 0.1 AU/mL for samples tested at 1:200 dilution and a >99% specificity cutoff of 0.3 AU/mL using a panel of pre-2020 samples (Figure. S1B). The anti-RBD IgG4 assay is more sensitive in comparison to anti-RBD total IgG, due to the lower background signals; in line with the much lower total IgG4 levels in sera in comparison to total IgG.

### Statistical analysis

Ratios of anti-RBD IgG4 titers divided by anti-RBD total IgG after third vaccination (V3) were analyzed using the Kruskal-Wallis test and the Conover-Iman post-hoc multiple comparisons test with Benjamini-Hochberg (FDR) correction. A lower limit of 0.005 was imputed for ratio values, as lower ratios only resulted from IgG4 titer values below seroconversion cutoff. Analysis and visualization was performed using R version 4.1.2 (39) with packages ‘tidyverse’ version 1.3.1 (40), ‘conover.test’ version 1.1.5, ‘scales’ version 1.2.1, ‘ggpubr’ version 0.4.0 and ‘patchwork’ version 1.1.2.

## Results

Samples from 604 participants of the ongoing Target-2-B! Immunity against SARS-CoV-2 vaccination cohort were retrieved for this study (Table 1). The mean age of participants was 53.0 years (SD 14.1) and 62.1% were female. Vaccinations were given between February 2021– May 2022; the median interval between first and second doses was 36 days (IQR 35 - 42) and 190 days (IQR 176 - 200) between second and third. Participants were sampled 28 days after each vaccination, and just prior to the third vaccination.

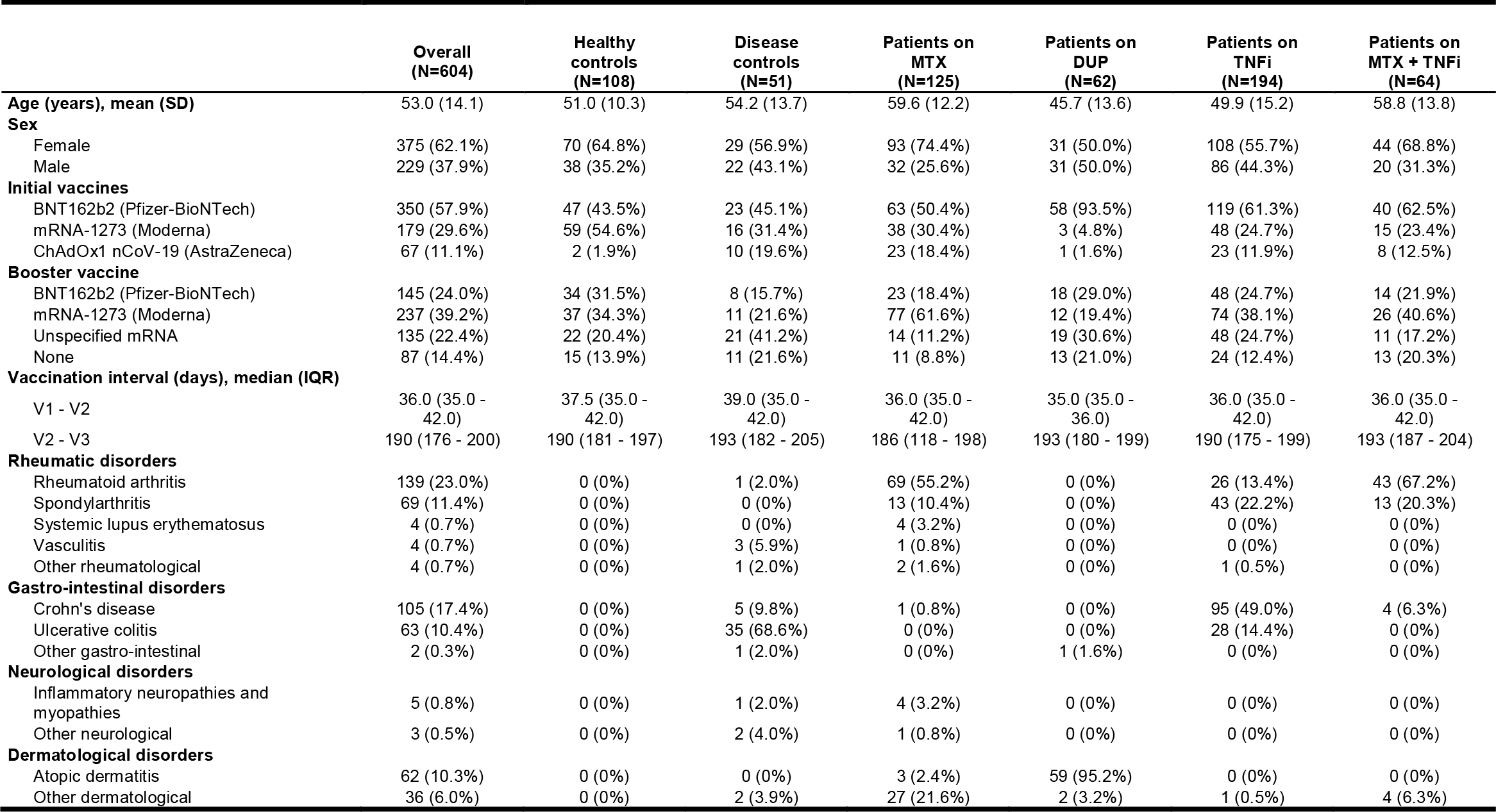

A robust anti-RBD total IgG response was observed at 28 days after the first and second mRNA vaccination dose. Antibody levels had declined by about an order of magnitude just prior to the third vaccination, and returned to post-second levels 28 days after the third dose in all groups (Figure 1A).

**Figure 1:**
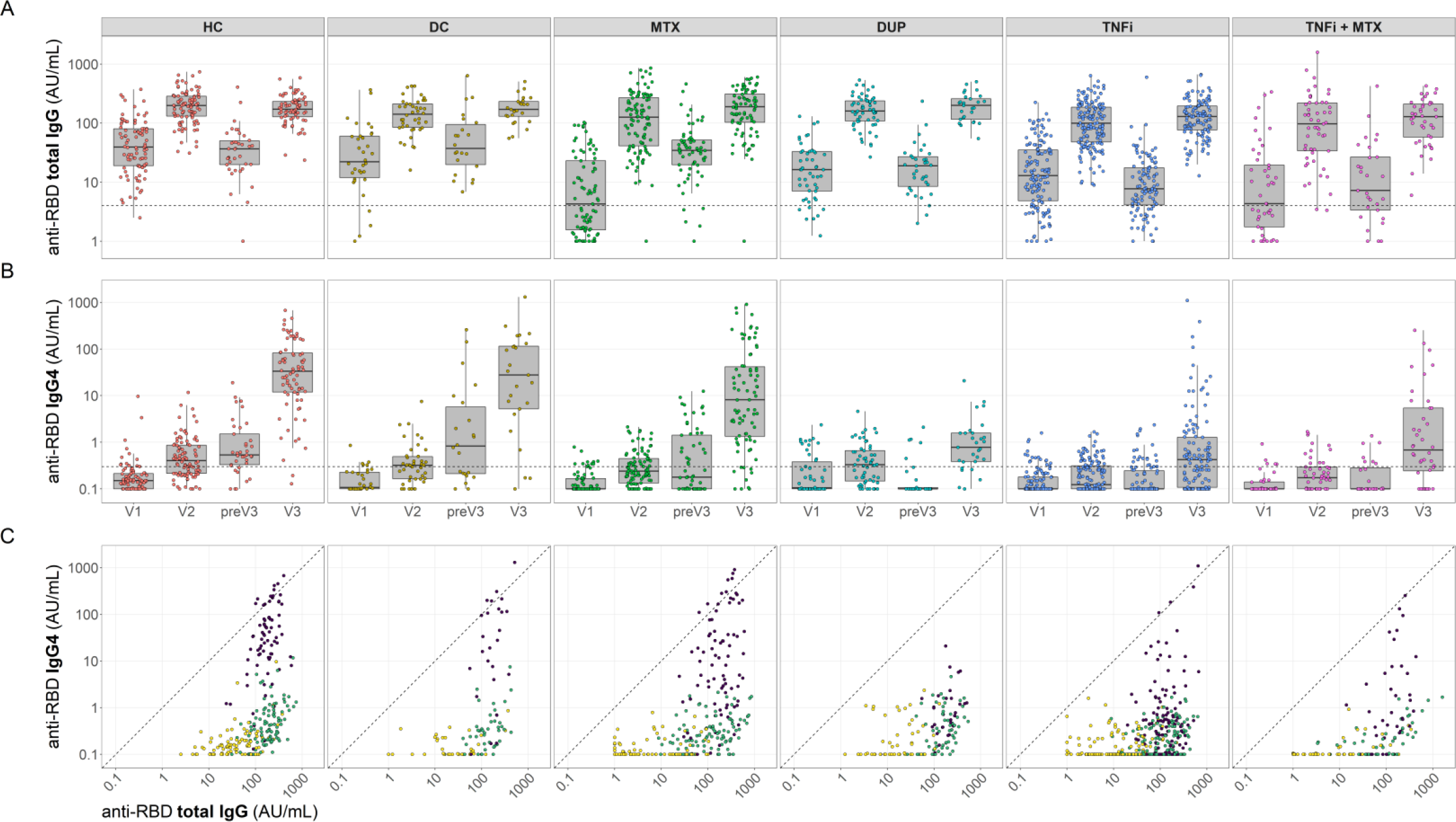
Longitudinal RBD-specific total IgG and IgG4 antibody response in healthy controls, disease controls and treatment groups after mRNA vaccination. Serum samples were collected 28 days after first, second and third vaccination (V1, V2, and V3) and immediately prior to third (preV3). Anti-RBD titers were assessed by direct ELISA and calculated in arbitrary units (AU) derived from pooled convalescent healthy donor plasma standards collected in early 2020 (total IgG) which was set at 100 AU/mL, or a monoclonal standard (IgG4) expressed in equivalent AU/mL. **A-B** Box plots showing RBD-specific total IgG (**A**) and IgG4 (**B**). Central lines in al box plots indicate the median, with hinges indicating 25^th^ and 75^th^ percentiles. Dashed lines represent seropositivity cutoffs, 4 AU/mL for total IgG and 0.3 AU/mL for IgG4, determined as the AU value where >99% of pre-pandemic samples were considered negative. **C** Scatter plots showing RBD-specific total IgG and IgG4, as in **A** and **B** respectively. Yellow dots indicate V1, green dots V2, and purple dots V3. Dashed diagonal lines indicate 1:1 titer ratio.

Anti-RBD IgG4 levels were very low after the first and second mRNA vaccination dose (Figure 1B, C). However, in HC and DC, the IgG4 antibody response was greatly boosted by the third mRNA vaccination dose (Figure 1B, C), yielding a sharp increase in the proportion of RBD-specific IgG4 (median 21.2% (IQR 6.6 – 44.7%) and 15.4% (IQR 2.2 – 78.8%) respectively; Figure 2). Conversely, IgG4 levels remained very low in patients treated with dupilumab compared to HC and DC, with a median proportion of less than 1%. TNFi treatment similarly reduced the induction of RBD-specific IgG4, both as single agent, as well as in combination with MTX. Patients treated with MTX monotherapy exhibited a modestly reduced induction of IgG4 skewing after the third dose with a median proportion of 6.7% (IQR 0.8 – 33.9%). Proportional induction of IgG4 was comparable across different combinations of BNT162b2 and mRNA-1273 as initial versus booster doses (not shown). In contrast, induction of IgG4 was virtually absent in all individuals initially vaccinated with two doses of ChAdOx1 nCoV-19, regardless of whether they were treated with immunosuppressive treatment (Figure S2). Taken together, we observe a profound suppression of repeated mRNA vaccination-driven RBD-specific IgG4 induction by both the IL-4R blocking antibody dupilumab and TNFi, while initial viral vector-based vaccination did not induce a relevant IgG4 response.

**Figure 2.**
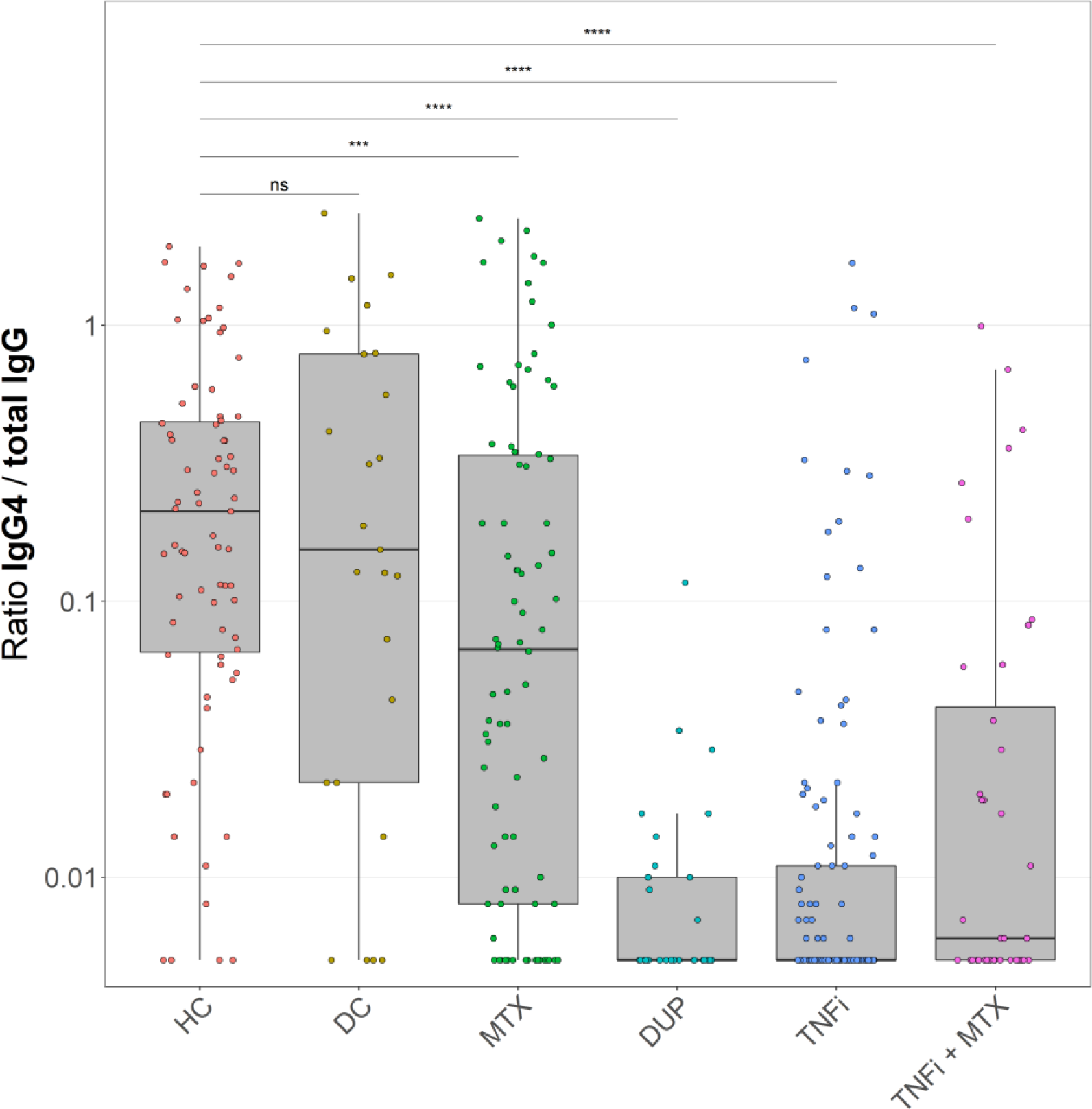
Ratio of RBD-specific IgG4 over RBD-specific total IgG in healthy controls, disease controls and treatment groups after third mRNA vaccination. Box plots showing ratios computed as RBD-specific IgG4 / RBD-specific total IgG in post-third vaccination samples (V3). In all box plots, central lines indicate the median, with hinges indicating 25^th^ and 75^th^ percentiles. Whiskers indicate the furthest data points up to 1.5 * IQR beyond hinges.

## Discussion

Here we longitudinally analyzed IgG4 RBD-specific antibody responses in a large cohort consisting of 604 participants overall. In accordance with previous findings we report an increase in RBD-specific IgG4 after third SARS-CoV-2 mRNA vaccination in HC and DC, which was virtually absent after primary adenoviral vector-based vaccination. The IgG4 skewing is profoundly reduced in patients treated with the IL-4R-blocking antibody dupilumab, as well as with widely used TNFi. Given the role of Th2 responses in IgG4 switching it may not be wholly unexpected that interference of IL-4 signaling impairs IgG4 induction. On the other hand, the inferred role for TNF signaling as evidenced by our results is surprising. These insights may further contribute to optimization of future mRNA vaccines and vaccination strategies, as well as tolerizing strategies, for instance for specific immunotherapy to relieve allergic symptoms.

Dupilumab blocks the IL-4/IL-13 receptor and is used to treat atopic dermatitis. These cytokines are typical for the Th2 axis that has been associated with IgG4 switching (25–31). Nevertheless, we are not aware of previous studies demonstrating inhibition of IgG4 skewing by dupilumab. T follicular helper (Tfh) cells are a major source of IL-4 and persistent Tfh cells were found after repeated SARS-CoV-2 mRNA vaccination, which could potentially facilitate IgG4 class switching (41). The suppressed RBD-specific IgG4 titer after third vaccination in the dupilumab-treated group indicates a pivotal role of IL-4 signaling in IgG4 switching, and may provide an opportunity for therapeutic intervention in case of undesired IgG4 responses. Furthermore, these findings may support the rationale behind the consideration of dupilumab as a potential treatment for IgG4-related disease (IgG4-RD), a group of immune-mediated fibrotic diseases that affect several organs and are characterized by high serum IgG4 levels plus increased tissue infiltrating IgG4^+^ plasma cells (42). The role that had been attributed to IL-4 together with Th2 cells in IgG4 class switching, led to several case studies treating IgG4-RD patients with dupilumab. However, controversial outcomes in these studies fuel a still ongoing discussion on whether dupilumab treatment is beneficial in IgG4-RD or not (43).

The pro-inflammatory cytokine TNF is the key driver of inflammation in many chronic inflammation settings and is secreted by various cell types, such as macrophages, T cells, B cells, and NK cells (44). Inhibition of TNF by a variety of TNFi suppressed the development of RBD-specific IgG4 after third mRNA vaccination. Total IgG anti-RBD levels were comparable to the healthy and untreated disease control groups, indicating a specific block in IgG4 switching in these patients. Interestingly, several TNFi (adalimumab in particular) have been extensively studied in light of anti-drug antibody development, a typical IgG4-skewed response. In these responses, adalimumab treatment has been shown to induce potent neutralizing anti-drug antibodies that shift towards IgG4 (45). It is somewhat paradoxical that substantial IgG4 skewing of these anti-adalimumab antibodies is observed under TNF blockade, while IgG4 skewing of the mRNA vaccination-induced immune response is markedly reduced. The extent of IgG4 switching may be dependent on a delicate balance between multiple pathways, whereby the nature of the antigen itself may be an important factor in directing this balance. Notably, total IgG4 serum levels are not affected by adalimumab treatment in general (25). We may conclude that a complete understanding of the mechanistic drivers responsible for IgG4 switching is still lacking.

Overall we have demonstrated significantly reduced IgG4 class switching by dupilumab as well as TNFi upon repeated mRNA vaccination for SARS-CoV-2, indicating a role for IL-4/IL-13 and TNF in IgG4 class switching in the context of mRNA vaccination.

## Data Availability

All data produced in the present work are contained in the manuscript

## Acknowledgements

We thank Julian Freen-van Heeren for stimulating discussions.

### Appendix T2B! Immunity against SARS-CoV-2 study group

Anneke J. van der Kooi, Joop Raaphorst, Mark Löwenberg, R. Bart Takkenberg, Geert R.A.M. D’Haens, Marc L. Hilhorst,, Yosta Vegting, Frederike J. Bemelman, Alexandre E. Voskuyl, Bo Broens, Agner R. Parra Sanchez, Cécile A.C.M. van Els, Jelle de Wit, Abraham Rutgers, Karina de Leeuw, Jan J.G.M. Verschuuren, Annabel M. Ruiter, Lotte van Ouwerkerk, Diane van der Woude, Renée C.F. van Allaart, Y.K. Onno Teng, Pieter van Paassen, Matthias H. Busch, Papay B.P. Jallah, Esther Brusse, Pieter A. van Doorn, Adája E. Baars, W. Ludo van der Pol, H. Stephan Goedee, Koos A.H. Zwinderman, Rivka de Jongh, Carolien E. van de Sandt, Olvi Cristianawati, Laura Fernandez Blanco, Niels J.M. Verstegen, Lisan H. Kuijper, Mariël C. Duurland, Ruth R. Hagen, Jet van den Dijssel, Christine Kreher, Amélie V. Bos, Virginia Palomares Cabeza, Veronique A.L. Konijn, George Elias, Marit J. van Gils, Elham S. Mirfazeli

