## Supplemental data for "Suppressed IgG4 class switching in dupilumab- and TNF inhibitor-treated patients after repeated SARS-CoV-2 mRNA vaccination"

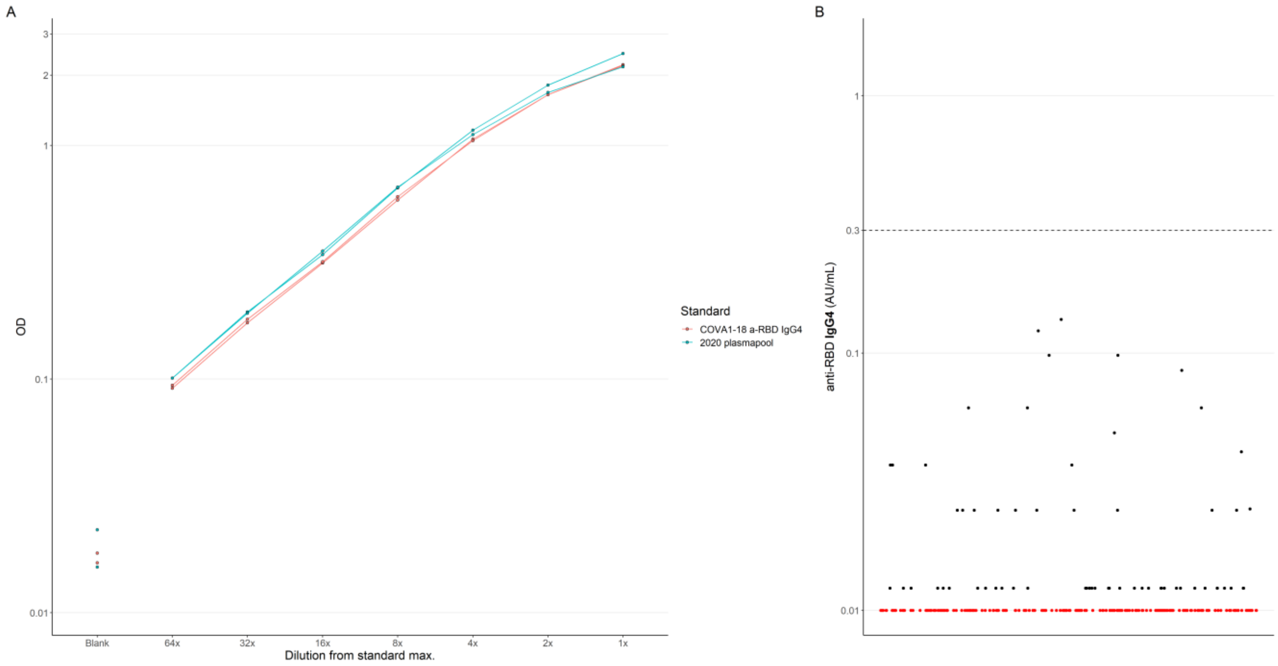


**Figure S1: validation of anti-RBD IgG4 ELISA. A** Line plot showing dilution curves of COVA1-18 anti-RBD IgG4 monoclonal and early 2020 plasmapool standard in anti-RBD total IgG ELISA across 2 representative experiments. **B** Scatter plot showing distribution of pre-2020 samples tested in anti-RBD IgG4 ELISA across 2 representative experiments, establishing 0.3 AU/mL cutoff for >99% specificity. Red data points indicate OD below technical limit of quantification; a titer value of 0.01 AU/mL was imputed for display purposes. As the range of measurements depicted here represent various levels of noise, 0.1 AU/mL was chosen as a meaningful lower limit of quantification.

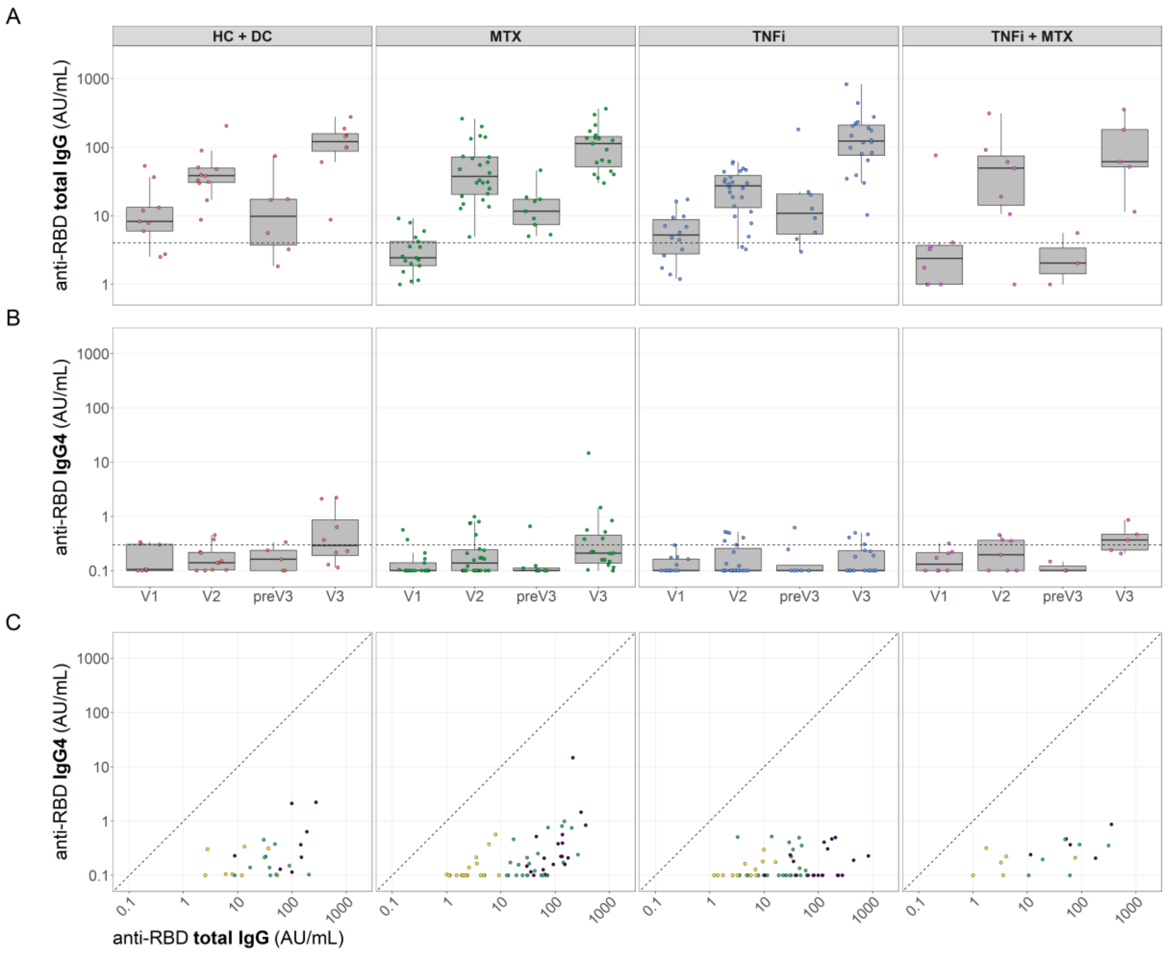


**Figure S2: RBD-specific total IgG and IgG4 antibody response in healthy controls, disease controls and treatment groups after primary adenoviral vector vaccination and mRNA boost. A-B** Box plots showing RBD-specific total IgG (**A**) and IgG4 (**B**). Dashed lines represent seropositivity cutoffs, 4 AU/mL for total IgG and 0.3 AU/mL for IgG4, determined as the AU value where >99% of pre-pandemic samples were considered negative. **C** Scatter plots showing RBD-specific total IgG and IgG4, as in **A** and **B** respectively. In all box plots, central lines indicate the median, with hinges indicating 25th and 75th percentiles. Whiskers indicate the furthest data points up to 1.5 * IQR beyond hinges.
